# Geometric Brain Signatures for Diagnosing Rare Hereditary Ataxias and Predicting Function

**DOI:** 10.64898/2026.03.08.26347882

**Authors:** Zuitian Tao, Gilles Naejie, Fuad Noman, Thiago J R Rezende, Marcondes Franca, Alex Fornito, Ian H Harding, Nellie Georgiou-Karistianis, Trang Cao, Susmita Saha, TRACK-FA Neuroimaging Consortium

## Abstract

**Background:** Hereditary cerebellar ataxias (HCAs) are rare neurodegenerative disorders characterised by progressive motor impairment and overlapping clinical phenotypes. Although genetic testing provides etiological diagnosis, diagnostic delays frequently arise before targeted testing, owing to non-specific presentation and limited clinician familiarity. Imaging-derived biomarkers that capture phenotypic expression and network-level consequences of disease could support earlier recognition of hereditary ataxia, guide appropriate genetic testing, and provide sensitive measures of disease evolution. Building on evidence that cortical geometry shapes functional organisation, we hypothesised that geometric signatures derived from structural magnetic resonance imaging (sMRI) could discriminate HCA subtypes and yield progression-sensitive biomarkers, while enabling scalable prediction of function.

**Methods:** We decomposed sMRI and task-evoked functional MRI data from three independent cohorts using cortical geometric eigenmodes, intrinsic spatial patterns defined by cortical surface geometry, to obtain structural and functional geometric signatures. Structural signatures were used to train neural networks for disease classification and to derive biomarkers sensitive to annual progression. We further modelled structure-to-function mappings to predict functional geometric signatures from sMRI and evaluated their diagnostic and longitudinal utility.

**Findings:** Our framework achieved high diagnostic performance, distinguishing healthy controls from Friedreich ataxia (FRDA) with a maximum AUC of 0.93 and separating FRDA from spinocerebellar ataxia type 1 (SCA1) and SCA3, with AUCs up to 0.81, showing cross-cohort generalisability. Structure-to-function-signature prediction achieved coefficient of determination up to 0.62 and correlation reaching 0.86 across health and disease, while predicted functional signatures improved classification beyond structural signatures alone and enabled partial reconstruction of individual task-activation map. Geometric brain signatures showed greater progression sensitivity than conventional volumetric MRI measures.

**Interpretation:** This geometry-driven framework offers novel, objective, multiscale biomarkers for diagnostic-decision-support and monitoring HCAs and provides proof-of-concept for the feasibility of predicting fMRI-equivalent biomarkers in disease from routine sMRI, which is far more practical in movement-disorder populations.

**Funding:** Friedreich Ataxia Research Alliance USA.

**Research in Context:** *Evidence before this study:* We searched PubMed, Scopus, IEEE Xplore, and Google Scholar, for peer-reviewed studies published up to 2025 using combinations of terms related to hereditary cerebellar ataxia (HCAs), Friedreich ataxia (FRDA), spinocerebellar ataxia (SCA), diagnosis, progression, MRI biomarkers, structural MRI (sMRI), functional MRI (fMRI), machine learning (ML), deep learning (DL), task activation maps, prediction, and geometric eigenmodes. We found that while sMRI studies in HCAs consistently showed patterns of cerebellar, brainstem, and supratentorial atrophy, highlighting their potential diagnostic value as non-invasive biomarkers, existing studies on imaging-based diagnostic tools for HCAs typically used small sample size, single-site data and focused on narrow classification tasks rather than generalisable frameworks for differential diagnosis. Clinically meaningful objective biomarkers sensitive to disease progression are also limited, with most outcome measures relying on subjective clinical rating scales or conventional MRI metrics with restricted sensitivity and reproducibility. In addition, fMRI reveals important network-level abnormalities in disease, however, motion artefacts, task-performance difficulties and long acquisition times limit its applicability in movement-disorder populations. Recent work in healthy populations showed that structural data could predict task-fMRI activation using DL, yet disease-specific and clinically deployable investigations remain unexplored. In parallel, advances in brain geometric eigenmode research underscore that cortical geometry provides a principled structural basis that shapes multi-scale functional organisation. However, no study has investigated cortical geometric signatures as tools to address three major challenges in hereditary ataxia research and clinical care: diagnostic delay, lack of progression-sensitive objective biomarkers, and practical limitations of functional imaging acquisition.

*Added value of this study:* Using a combined geometric and ML framework, we showed that cortical geometric signatures captured multiscale brain organisation that constitute novel, generalisable biomarkers for differential diagnosis across HCA subtypes. Our models reliably distinguished healthy controls from individuals with FRDA, demonstrated consistent performance across independent cohorts, and further separated FRDA from multiple SCA subtypes. Importantly, we provided proof-of-concept for structure-to-function prediction and showed that fMRI-equivalent functional signatures could be inferred from sMRI in both health and disease, enabling reliable approximation of individual task-activation maps without requiring fMRI acquisition. Incorporating these predicted functional signatures improved diagnostic accuracy beyond structural measures alone. Both structural and predicted functional biomarkers demonstrated greater sensitivity to annual disease progression than conventional volumetric metrics, with comparable performance to clinical scales.

*Implications of all the available evidence:* Cortical geometric signatures pave the way for a clinically deployable neuroimaging diagnostic decision-support tool that could guide clinicians toward targeted genetic testing and potentially reduce diagnostic delay in HCAs. These biomarkers provide objective, rater-independent measures of disease evolution that are more scalable and reproducible than clinical ratings alone, and more sensitive than conventional imaging measures, with important implications for improving clinical trial design and monitoring therapeutic efficacy. The ability to infer functional signatures from sMRI allows clinicians to probe aspects of functional organisation and network disruption using routine sMRI, which is substantially more practical in movement-disorder populations, thereby introducing a novel fMRI-equivalent biomarker that may further improve diagnosis and progression monitoring.

## Introduction

Hereditary cerebellar ataxias (HCAs) are rare, progressive neurodegenerative movement disorders characterised by heterogeneous motor and non-motor symptoms, wide variability in age of onset, and diverse trajectories of functional decline.^1–4^ Although current genetic diagnostics, particularly repeat-expansion assays and targeted panels, provide efficient and reliable etiological confirmation for common HCAs, patients frequently experience prolonged diagnostic pathways. In clinical practice, these delays arise from phenotypic overlap between ataxia symptoms, variable referral pathways, limited clinician familiarity and limited genetic test access.^5–9^ In this context, tools that support early phenotypic characterisation may help clinicians recognise when a particular HCA subtype is likely, guide timely and targeted genetic investigations, and ultimately shorten the path to definitive diagnosis.

Even after genetic diagnosis is established, major bottlenecks remain in monitoring disease evolution.^10–13^ HCAs are increasingly understood as disorders of distributed cerebro-cerebellar systems, in which primary pathology in the cerebellum and its afferent/efferent connections can have widespread downstream consequences for cortical organisation and large-scale networks.^14^ This systems-level perspective motivates the search for biomarkers that capture phenotypic expression and network-level consequences of disease signals that are not available from genotype alone.

Current outcome assessment relies heavily on clinical rating scales. The Scale for the Assessment and Rating of Ataxia (SARA) and the modified Friedreich Ataxia Rating Scale (mFARS) remain central endpoints in natural history studies and clinical trials because they are sensitive to clinically meaningful change. However, these measures are inherently subjective and rater-dependent. They may show non-linear sensitivity across disease stages (e.g., reduced responsiveness in non-ambulatory FRDA patients) and provide only coarse summaries of complex neurobiological processes.^1,11,13,15–17^ On the other hand, recent work highlights the value of structural magnetic resonance imaging (sMRI) to extract objective biomarkers for diagnosis and progression tracking in HCAs.^13^ SMRI consistently demonstrates progressive cerebellar and brainstem atrophy across HCA subtypes, with distinct and variable involvement of cerebral regions. For example, spinocerebellar ataxia type 1 (SCA1) is characterised by early pontine and cerebellar degeneration frequently accompanied by cortical thinning and frontal and parietal atrophy;^10,18^ SCA3 shows widespread cerebellar and brainstem involvement with milder supratentorial changes emerging later in the disease course;^19^ and FRDA follows a distinct pattern marked by dentate nucleus atrophy, spinal cord thinning, and degeneration of the superior cerebellar peduncles and corticospinal tracts, with limited late-stage supratentorial involvement.^20,21^ Although such signatures inform clinical reasoning, automated and generalisable imaging models for differential diagnosis in HCAs remain scarce.^22–24^ Existing studies are frequently single-site, underpowered, focused on narrow classifications, or reliant on local voxel/vertex/ROI analyses that can be sensitive to analytic choices and reduce reproducibility across cohorts.^25^ In addition, conventional sMRI measures have shown limited sensitivity to longitudinal changes.^26,27^ Objective, reproducible biomarkers derived from a widely available imaging modality (e.g. sMRI) that enable earlier diagnostic decision support and more sensitive progression monitoring are therefore urgently needed.

Functional MRI (fMRI) has provided important insights into network-level dysfunction in FRDA and SCAs, including altered cortical activation and connectivity patterns that complement structural findings.^28,29^ However, fMRI remains challenging to acquire in movement-disorder populations because of motion artefacts, task-compliance demands, and variability, which has limited both research use and clinical translation.^13,30^ Approaches capable of inferring functional signatures from more accessible modalities such as sMRI could help overcome these barriers. To date, only one deep-learning (DL) study has attempted structure-to-function prediction in healthy individuals, but it reported only indirect evidence of individual-level correspondence (comparison of self-vs-other correlations) without providing any quantitative accuracy of predicted task-activation maps.^31^ Disease-specific investigations, therefore remain lacking.

To address these clinical and translational challenges in HCAs, we sought an imaging representation that is anatomically grounded yet sensitive to multiscale network organisation, interpretable, scalable, and reproducible across sites, and naturally suited to linking structure and function. Cortical geometric eigenmodes provide such a representation. They are intrinsic spatial patterns determined by the geometry of the cortical surface, and their ordered, multiscale structure, from coarse, low-frequency modes to fine, high-frequency modes, allows simultaneous sensitivity to widespread network abnormalities and more localised cortical changes.^32^ Recent work shows that any individual cortical map, whether structural or functional, can be expressed as a weighted combination of these modes, each with a characteristic spatial fingerprint.^33–35^ This enhances reproducibility and reduces dependence on analytic choices, while the associated mode weights serve as personalised structural or functional geometric signatures. Finally, because structural and functional maps share the same geometric constraint, eigenmodes provide a natural bridge between anatomy and function.^33,34^

In this study, we developed and validated a geometry-driven DL framework that used cortical eigenmodes to (i) derive multiscale, interpretable sMRI biomarkers and evaluate their performance for differential classification across major HCA subtypes (with FRDA as the primary case study and comparisons including SCA1 and SCA3); (ii) model structure-to-function mappings to predict fMRI-equivalent functional signatures from sMRI alone, providing scalable access to functional signatures when fMRI is unavailable; and (iii) assess the sensitivity of structural and predicted functional geometric signatures to longitudinal change, benchmarking against clinical and volumetric MRI measures.

## Methods

We developed diagnosis and prediction models by integrating eigenmode decomposition with DL approaches and using sMRI and task-fMRI data from participants across three independent cohorts: the Belgium dataset,^36^ the TRACK-FA dataset,^37^ and the Campinas dataset^38–40^ (see Supplementary Tables S1, S2, and S3 for demographic details). Figure 1 presents an overview of the workflow. Detailed descriptions of participants, MRI acquisition protocols, preprocessing, eigenmode derivations, feature selection, model architectures, training, evaluation and performance analysis^41^ are provided in the Supplementary Methods. Although HCAs are primarily cerebellar disorders, we analysed only cortical data in this study for three reasons. First, as described above, HCAs are now well-understood to affect distributed cerebro-cerebellar networks, with established evidence of diaschisis and secondary cortical reorganisation that extends beyond the cerebellum and brainstem.^20,28,29,42^ Second, cortical geometric eigenmodes can be reliably generated using established pipelines.^33,34^ Unlike the thin-sheet neocortex, the cerebellum’s complex three-dimensional structure requires volume-based rather than surface-based eigenmode estimation. Such volume-based approaches have been demonstrated for subcortical structures in work by Pang and colleagues,^34^ but have not yet been systematically investigated on the cerebellum. Establishing cerebellum-specific pipelines, therefore remains an important direction for future work. Third, cortical geometric eigenmodes are derived from population-averaged cortical surface, providing a common basis that do not suffer from processing or site-specific bias,^25^ thereby enabling cross-cohort consistent analysis for robust diagnostic and longitudinal applications.

**Figure 1.**
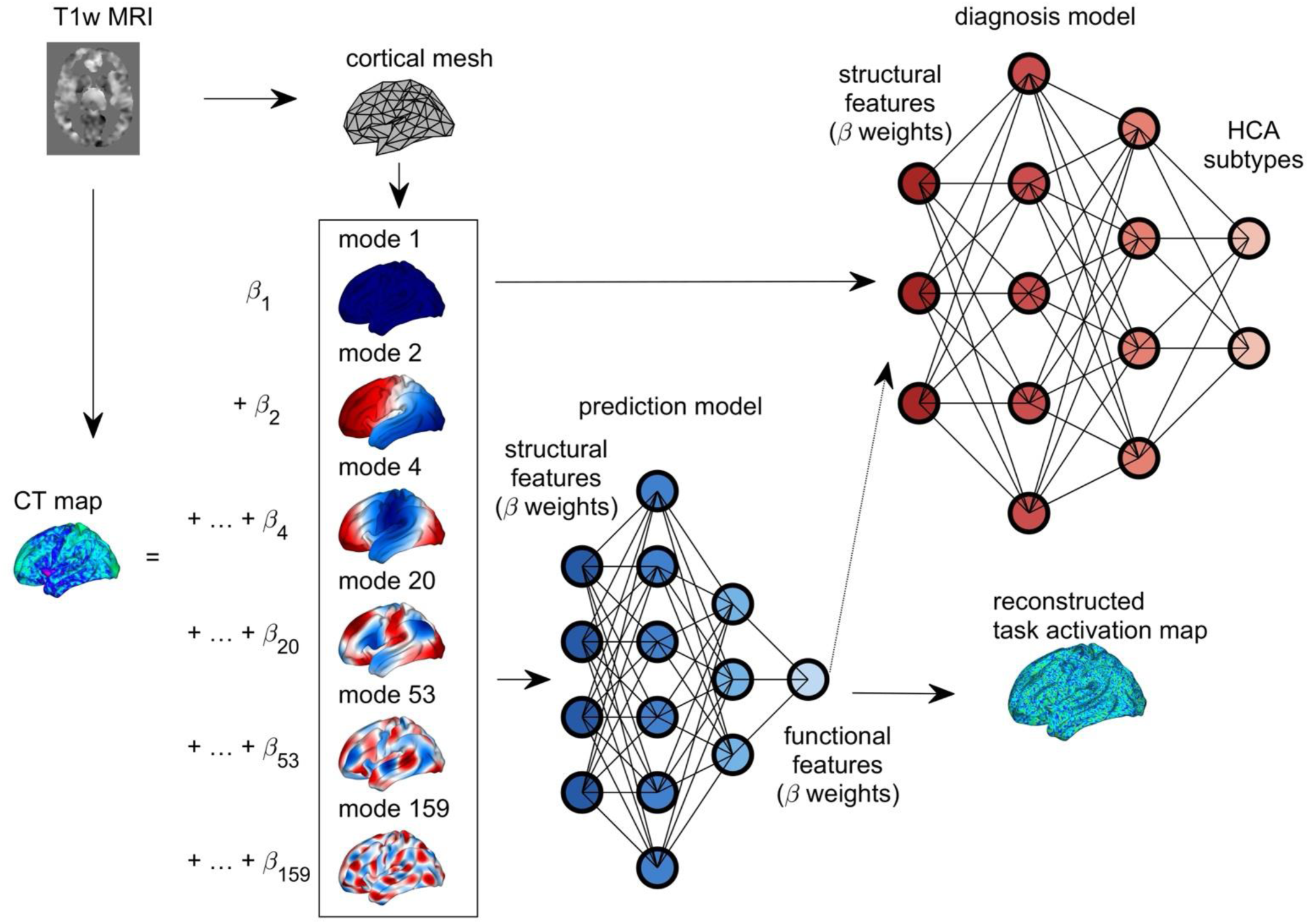
Overview of the geometry-driven DL framework linking cortical structure to functional activation and diagnosis. A set of geometric eigenmodes are derived from the population-averaged cortical surface mesh obtained from T1-weighted (T1w) MRI. Individual cortical thickness maps extracted from T1w MRI are decomposed as weighted linear combinations of these modes (β₁…β200) and the resulting eigenmode weights constitute the structural features. Task-evoked fMRI activation maps from the passive-movement task are decomposed in the same way, with their eigenmode weights serving as functional features. Selected structural features are input into a neural-network prediction model to predict functional features, which are then used to reconstruct individual task-activation maps. In parallel, selected structural, empirical functional and/or predicted functional features are provided to neural network diagnosis models to generate diagnostic labels (HCA subtypes).

### HCA Diagnosis With Structural Geometric Features

To develop the HCA diagnosis model, cortical thickness maps of each hemisphere, derived from T1-weighted MRIs, from the Belgium dataset (21 healthy controls (HCs), 20 FRDA) were decomposed using 200 geometric eigenmodes to obtain the β-weight of each mode. As the brain is asymmetric,^43^ our models consider each hemisphere separately in order to identify hemisphere-specific signatures. Feature selection and evaluation of the DL diagnosis model were nested within a leave-one-out cross-validation framework. Within each training fold, mode-specific β-weights were compared between groups using two-sample *t*-tests (uncorrected *p* < 0.05). Justification for the use of uncorrected statistical significance is provided in Supplementary Methods 4. Significantly different β-weights, i.e., structural geometric signatures, constitute the structural input features into a fully connected neural-network classifier for FRDA versus HC diagnosis (Supplementary Methods 5.1). Cross-cohort generalisability was evaluated via transfer learning in the larger, independent, multi-site TRACK-FA cohort (n=168 FRDA; n=92 HCs) (Supplementary Methods 5.3). For differential diagnosis among HCA subtypes, separate models were trained to classify FRDA vs SCA1, FRDA vs SCA3, and SCA1 vs SCA3 using the Campinas dataset, with five-fold cross-validation. We then benchmarked our models using geometric features with a model using conventional features derived from principal component analysis (PCA) of the CT maps from Belgium dataset (Supplementary Methods 6). Model performance was evaluated using AUC, accuracy, precision, recall, and F1-score, with AUC considered as the primary metric for model comparison.

### HCA Diagnosis With Functional Geometric Features

In addition to deriving HCA diagnosis from structural MRI, we hypothesised that task-based fMRI could further improve diagnostic performance. Accordingly, cortical functional activation maps from passive-movement-evoked fMRI in the Belgium dataset were decomposed using geometric eigenmodes to obtain mode β-weights. Functional features were selected from statistical test similar to structural features and used either alone or in combination with structural features for the diagnostic models, following the same training and cross-validation strategy as in the structural models described above (Supplementary Methods 5.1). A similar benchmarking strategy was applied using PCA-based models, consistent with the structural approach.

### Prediction Of Functional Geometric Features Solely From Structural Geometric Features

Because fMRI acquisition is often impractical in ataxia cohorts and limits the availability of functional biomarkers, we trained neural network regression models to predict functional geometric signatures, i.e., β-weights of fMRI task activation maps, solely from structural geometric signatures, i.e., β-weights of cortical thickness maps. We used both HCs and FRDA patients’ data from the Belgium cohort. Model training, feature selection, and evaluation procedures are detailed in Supplementary Methods 5.2. Predicted functional β-weights were also used to reconstruct task-activation maps, with reconstruction accuracy quantified as the correlation between empirical and reconstructed maps (Supplementary Methods 3.2). PCA-based models were again used for benchmarking.

### Diagnostic Utility Of Predicted Functional Geometric Features

The structure-to-function prediction models trained on the Belgium dataset were applied to TRACK-FA structural MRI to generate predicted functional features (Supplementary Methods 5.2). These predicted features were then used to fine-tune the Belgium-trained diagnostic models, whether originally based on functional features alone or on combined structural-and-functional features, via transfer learning, allowing us to evaluate the added diagnostic utility and cross-cohort robustness of the predicted functional signatures.

### Sensitivity of Geometric Features to Disease Progression

We quantified disease-progression sensitivity in TRACK-FA by calculating effect sizes (Cohen’s *d)* for each structural and functional geometric feature between baseline and 1-year follow-up (Supplementary Methods 7). Features were considered sensitive if the longitudinal changes met these criteria: (i) exhibiting opposite directions of change between the HCs and FRDA groups, (ii) between-group difference in effect size exceeding a manually defined threshold (|*Δd*|> 0.3) and (iii) statistically significant (two-sample *t*-tests, *p* < 0.05; uncorrected) (Supplementary Methods 7).

### Linking Eigenmodes to Functional Networks

To improve interpretability and relate geometric biomarkers to known functional brain systems, we characterised the functional network signatures of each eigenmode. Mean absolute amplitudes were computed within cortical functional networks defined by the Yeo 7-network atlas^44^ in each eigenmode and the network exhibiting the highest amplitude was defined as the dominant one for that eigenmode. This analysis allowed us to map diagnostically and progression sensitive eigenmodes onto established functional systems, providing a neurobiological interpretation of the multiscale patterns identified by the models (Supplementary Methods 8).

## Results

### HCA Diagnosis With Structural Geometric Features

We analysed structural geometric features, i.e., β-weights quantifying how strongly individual’s CT map expresses the spatial pattern of each eigenmode. Using this approach, we identified several eigenmodes showing significant group differences (two-sample t-test, p < 0.05) in β-weights between HCs and FRDA patients in the Belgium cohort (Figure 2a and b). These findings indicate both widespread large-scale disruptions and more spatially localised changes in FRDA, spanning across three canonical frequency bands: low (modes 1-50, wavelength ∼60 mm)), mid (51-100, ∼40 mm), and high-frequency (101-200, ∼30 mm). Modes with significant β-differences are listed in Supplementary Tables S4. Note that modes 2, 4, 20, 41, 53, and 159 showed significant β-differences in both hemispheres, suggesting consistent bilateral alterations in thickness patterns. Mapping the spatial expression of these modes onto canonical functional networks revealed that the dominant lobes of the altered eigenmodes preferentially overlapped limbic, somatomotor, and ventral attention/salience and frontoparietal control regions. This pattern suggests that cortical territories aligned with these networks are preferentially involved in the thickness alterations observed in FRDA.

**Figure 2.**
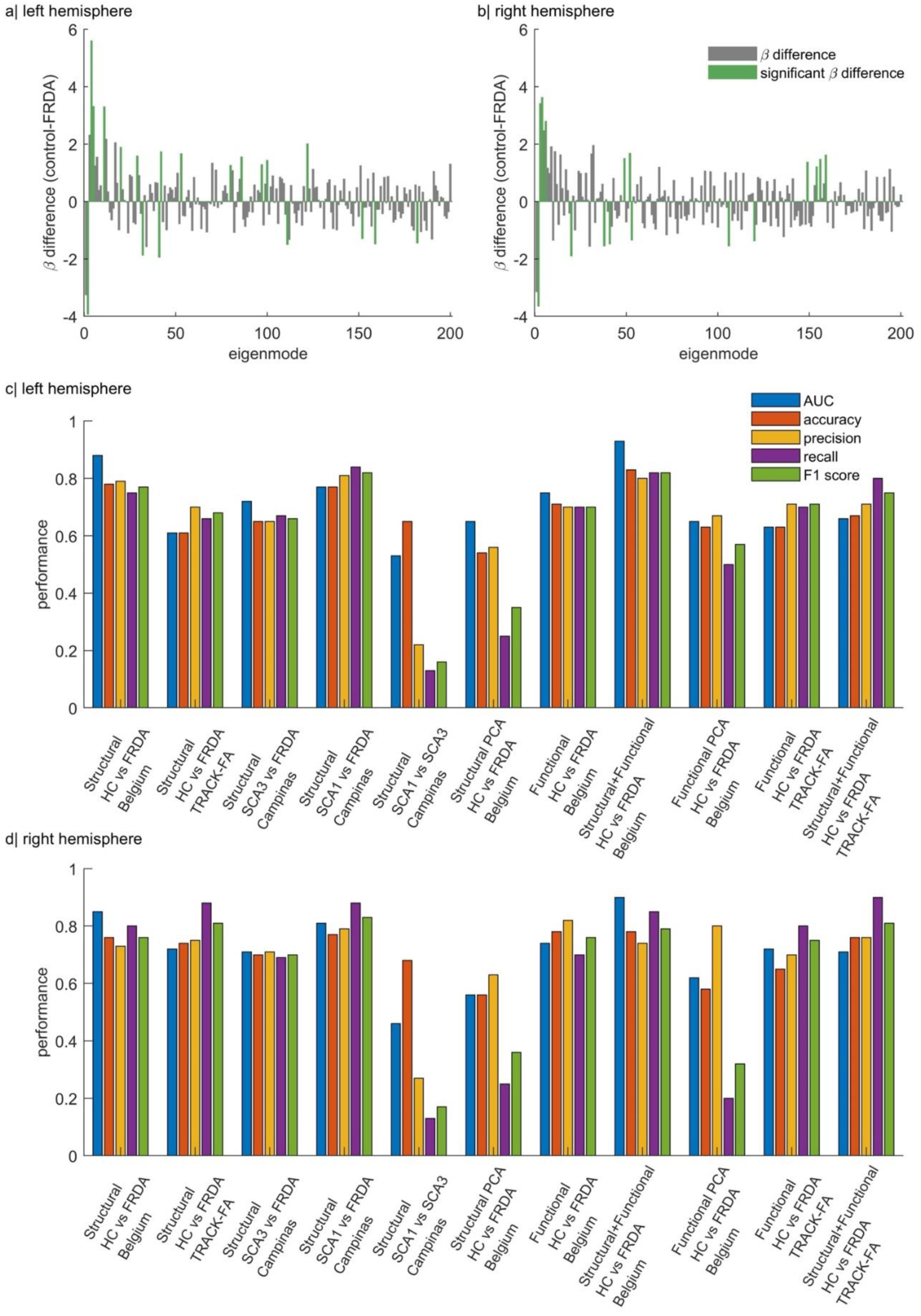
Illustration of diagnostic features and model performance. Differences in structural eigenmode weights between HCs and FRDA patients are shown for eigenmodes 1–200 in the a| left hemisphere and b| right hemisphere. Performance of diagnostic models using c| left hemisphere features and d| right hemisphere features is quantified using AUC, accuracy, precision, recall, and F1 score. Note that in the Belgium dataset, functional denotes features derived from empirical fMRI data, whereas in TRACK-FA, it refers to predicted functional features.

Comprehensive diagnostic performance measures for all models are provided in Supplementary Tables S5-S6 and Figure 2c–d. The HC-FRDA diagnostic model trained on structural geometric features achieved strong performance in both hemispheres, e.g., AUC of 0.88 and 0.85 in the left and right hemispheres, respectively. The transfer-learning model using structural geometric features from the TRACK-FA dataset showed similar performance to the Belgium-trained model, with a slightly higher performance in the right hemisphere compared to the left.

In differential diagnosis among HCA subtypes, classification performance varied across subtype comparisons. In particular, models distinguishing SCA1 vs FRDA achieved the highest performance (AUC: 0.81, right and AUC: 0.77, left) while the SCA1 vs SCA3 models exhibited the lowest (AUC: 0.53, left). Overall, these findings suggest that the weights associated with geometry-constrained, multiscale cortical thickness patterns constitute promising and potentially generalisable biomarkers for automated differential diagnosis of HCAs. However, distinguishing between closely related SCA subtypes remains challenging. The modes exhibiting significant β-differences for these models are listed in Supplementary Tables S7-9. These modes span both large-scale and localised patterns. In the FRDA versus SCA1 differentiating, modes 1, 4, and 9 showed bilateral significance, whereas in FRDA versus SCA3, modes 1, 4, and 108 were bilaterally altered. Spatially, dominant lobes of these modes align preferentially with limbic, somatomotor, and frontoparietal control territories in FRDA vs SCA1 differentiating, and with limbic and somatomotor territories in the FRDA vs SCA3 differentiating. In benchmarking against geometric features, conventional structural PCA-based features derived from the Belgium dataset showed markedly poorer performance in HC-FRDA diagnosis.

### HCA Diagnosis With Functional Geometric Features

Consistent with the structural findings, uncorrected significant group differences in eigenmode β weights derived from task-activation maps in Belgium data were observed in both hemispheres (Supplementary Tables S10 and Figures S1). Modes 1, 4, 63, 99, and 122 exhibited significant β weight differences in both hemispheres. Spatial mapping analysis showed that the dominant regions expressing these altered modes overlapped primarily with limbic, somatomotor, dorsal and ventral attention, default mode, and visual networks. The diagnostic models trained using functional geometric features achieved performance (AUC: 0.75, left) comparable to those based on structural features. Combining structural and functional geometric features from the Belgium dataset resulted in strong diagnostic performance (AUC: 0.93, left), surpassing that of single-modality models. These results suggest that geometry-driven structural and functional features capture complementary neuroimaging signatures of disease. Moreover, benchmarking against diagnostic models using functional PCA-based features revealed markedly inferior classification performance relative to functional geometric models.

### Prediction Of Functional Geometric Features Solely From Structural Geometric Features

To examine whether fMRI-equivalent features can be reliably predicted from sMRI alone, thereby providing a practical alternative when fMRI is unavailable, we trained 200 separate models, each predicting one functional geometric feature from the corresponding set of structural geometric features. Across all models, prediction performance reached a mean coefficient of determination (R²) of 0.26 ± 0.22 (range: −0.70 - 0.56) with a mean correlation of 0.68 ± 0.10 (range: 0.24 - 0.86) in the left hemisphere and R² of 0.24 ± 0.17 (−0.51 - 0.62) with a correlation of 0.63 ± 0.12 (range: 0.25 - 0.86) in the right hemisphere. Although the average explained variance was modest, the wide range of metrics indicates that specific functional features were predicted with relatively high accuracy, highlighting that structural MRI captures substantial information about particular aspects of functional organisation in both HCs and FRDA patients. Functional geometric features that achieved high predictive performance (R² > 0.5 and correlation > 0.7) are listed in Supplementary Tables S11. To further characterise the structural basis of these highly predictable modes, we examined the distribution of their top 25 structurally correlated modes across low, mid and high frequency bands as defined above. Band Contribution Ratio, defined as the number of correlated modes within a band divided by the number of structural modes available in that band, controls for bandwidth differences and allows direct comparison across frequency scales. In the right hemisphere (Supplementary Tables S13), controls showed slightly higher contributions of low-frequency modes to the predicted features (0.140) compared with the mid- (0.122) and high-frequency bands (0.119), whereas in FRDA patients a relative shift of the contributions towards the mid-frequency band was observed (Low: 0.105, Mid: 0.145, High: 0.125). However, left hemisphere structure to function mapping patterns were broadly preserved in health and disease.

Using 200 eigenmode weights derived from empirical task-activation maps, mean reconstruction accuracy across all subjects approached 0.6 in both hemispheres (range of correlations across individuals: 0.45-0.79 for the left hemisphere, 0.46-0.77 for the right), indicating a moderate-to-strong correspondence between the reconstructed and empirical activation maps (Figure 3). This level of accuracy is comparable to prior work,^34^ which reported >80% accuracy when using group-averaged rather than individual-level activation maps, as examined here. With predicted functional eigenmode weights, reconstruction accuracy remained moderate, with mean correlations of 0.39 (range: 0.19 - 0.60) in the left hemisphere and 0.36 (range: 0.08 - 0.65) in the right hemisphere across all subjects, with slightly higher performance observed when only HCs are analysed (see values for different cohorts in Supplementary Tables S12). These findings highlight two key points: (i) eigenmode decomposition captures a substantial proportion of task-related variance in functional activation, and (ii) functional features predicted from structural features preserve meaningful information about individual brain activity patterns.

**Figure 3.**
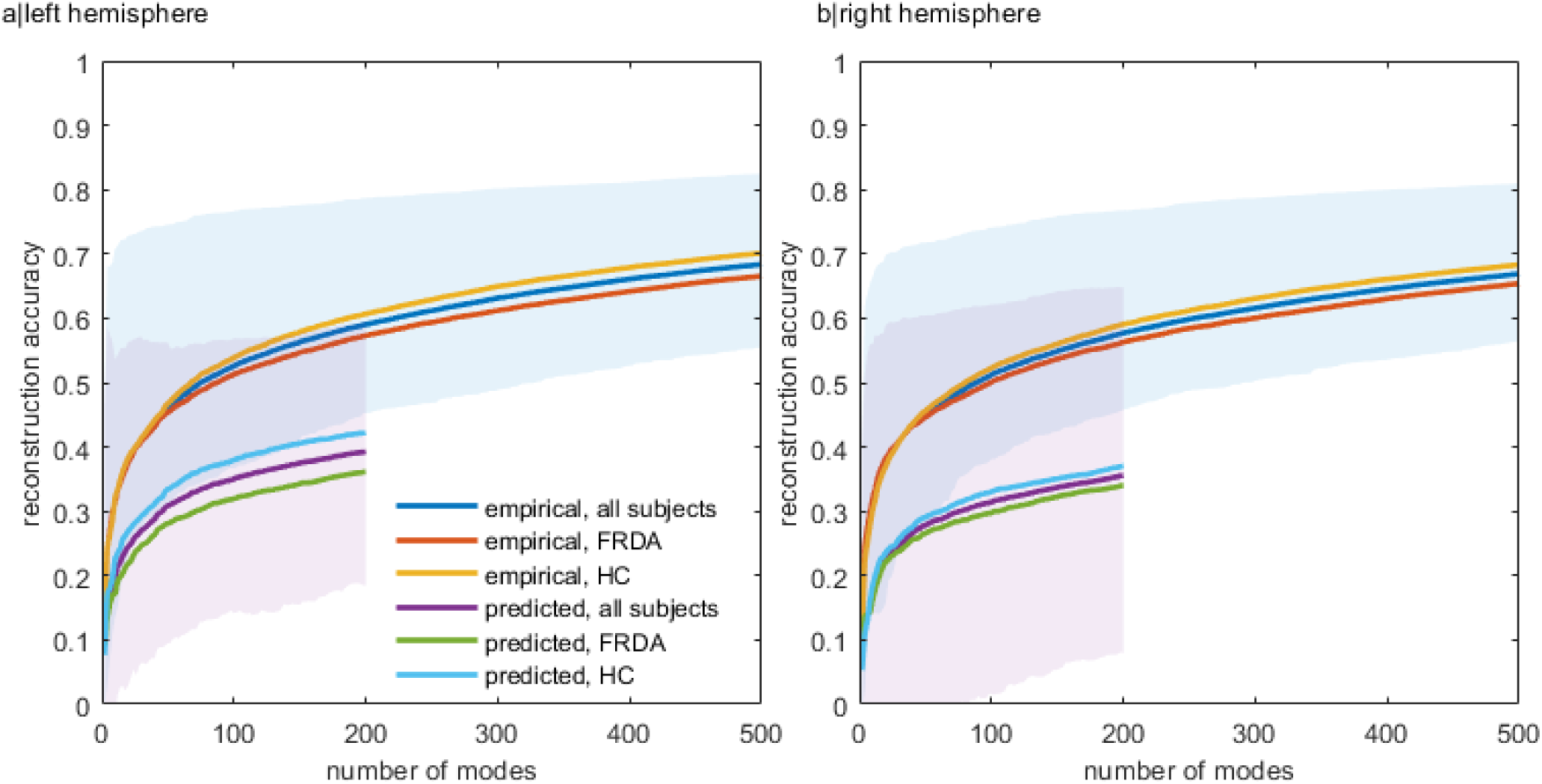
Reconstruction accuracy of task-activation maps using cortical eigenmodes. Mean correlation between empirical and reconstructed individual task-activation maps in the Belgium dataset, using weights derived directly from empirical activation maps or predicted from structural features, as a function of the number of cortical eigenmodes for the a| left and b| right hemisphere. Results are shown separately for HCs, FRDA patients, and all subjects. Shaded regions indicate the range of reconstruction accuracy across individuals.

In benchmarking analyses, PCA-based models achieved a mean R² of −0.19 ± 0.18 (range: −1.10 to 0.16) and a correlation of −0.00 ± 0.20 (−0.46 to 0.51) in the left hemisphere, and a mean R² of −0.15 ± 0.19 (−1.69 to 0.20) with a correlation of −0.03 ± 0.20 (−0.45 to 0.49) in the right hemisphere. These near-zero or negative R² values and negligible correlations indicate minimal predictive capacity, underscoring that geometry-informed representations capture structure–function relationships far more effectively than conventional PCA-based dimensionality reduction.

### Diagnostic Utility Of Predicted Functional Geometric Features

Functional geometric features were predicted by applying the Belgium-trained structure-to-function model to the large, independent TRACK-FA cohort, which did not include fMRI data (Supplementary Methods 5.3). To evaluate the clinical utility of these predicted functional features, we applied the diagnostic model trained on empirical functional geometric features from the Belgium dataset to the predicted functional features in TRACK-FA using a transfer-learning framework (Supplementary Methods 5.3). The model achieved moderate diagnostic performance (AUC: 0.72, right), comparable to that obtained using empirical functional features, particularly in the right hemisphere (Supplementary Tables S5–S6; Figure 2c-d). When structural and predicted functional geometric features were combined in the TRACK-FA cohort, diagnostic performance further improved relative to models using either modality alone and approached the accuracy achieved when combining empirical structural and functional features in the Belgium dataset, albeit with a lower AUC (Supplementary Tables S5-6; Figure 2c-d). These findings demonstrate the strong potential of a structure-to-function prediction approach for diagnostic applications.

### Sensitivity Of Geometric Features To Disease Progression

We next examined the sensitivity of geometric features to longitudinal disease progression in the TRACK-FA dataset over a 1-year interval, using Cohen’s *d* as an effect-size measure. Several structural geometric features, eigenmodes 23, 52, 57, 100, 116, 119, 144, and 179 in the left hemisphere (Figure 4a) and eigenmodes 40, 69, 143, 153, 168, and 184 in the right hemisphere (Figure 4b), met our biomarker criteria (Supplementary Methods 7). These findings indicate that annual disease progression in FRDA predominantly affects mid- to high frequency eigenmodes, consistent with increasingly regional and localised structural alterations over time. This pattern contrasts with the HC-FRDA diagnostic model, in which both low- and high-frequency modes contributed to classification. Thus, while global-scale abnormalities define the disease signature at baseline, longitudinal progression appears to be driven by increasingly local disruptions, i.e., lower spatial scales. Spatial mapping of the dominant lobes of these progression-sensitive structural modes showed preferential alignment with cortical territories spanning ventral attention/salience, default mode, visual, limbic, dorsal attention, and somatomotor networks, consistent with distributed cortical progression embedded within large-scale functional architectures.

**Figure 4.**
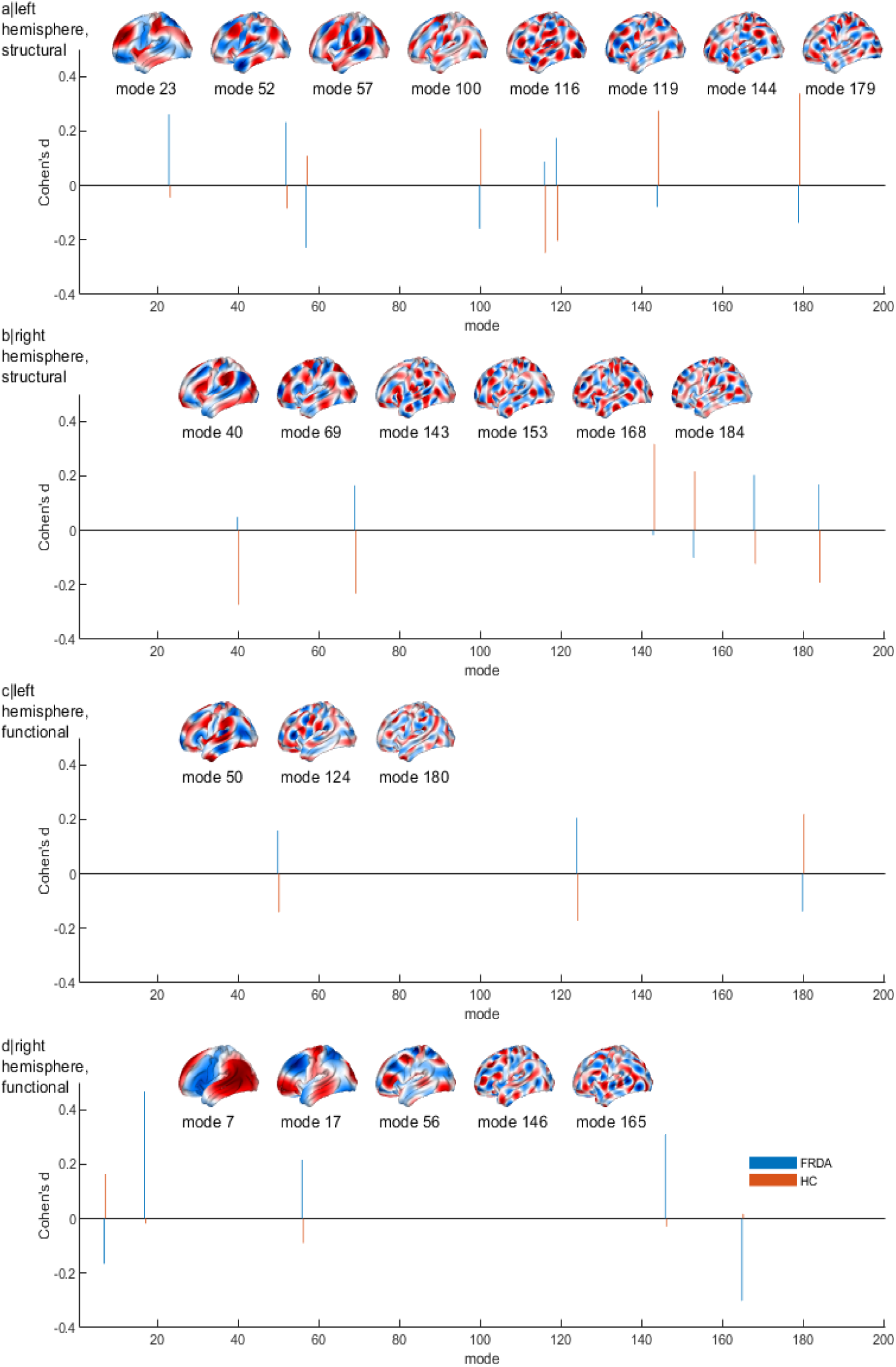
Geometric features sensitive to annual disease progression. Geometric features showing opposite directional change over time, with absolute between-group differences in Cohen’s *d* greater than 0.3 (*Δd* > 0.3) and showing statistically significant (two-sample *t*-tests, *p* < 0.05; uncorrected) between-group differences, are presented alongside their corresponding *d* values in HCs and FRDA patients. Panels a| and b| show structural geometric features in the left and right hemispheres, respectively, while panels c| and d| show predicted functional geometric features. Blue and orange vertical lines denote *d* values for HCs and FRDA patients, respectively.

With predicted functional features, the sensitive modes were eigenmodes 50, 124, and 180 in the left hemisphere (Figure 4c) and eigenmodes 7, 17, 56, 146, and 165 in the right hemisphere (Figure 4d). These findings, together with spatial mapping, suggest hemispheric asymmetry in functional progression. In the left hemisphere, the dominant lobes of progression-sensitive modes aligned primarily with limbic and dorsal attention territories, indicating localised disruption within these network-aligned regions. In contrast, right-hemisphere progression-sensitive modes reflected disruption of both lower-frequency, more global functional organisation and higher-frequency, localised components, with dominant involvement of default mode, dorsal and ventral attention, and frontoparietal control territories. In the left hemisphere, structural features generally exhibited greater sensitivity to progression than functional features, with structural eigenmode 23 showing the highest sensitivity (*d* in FRDA = 0.262). Conversely, in the right hemisphere, functional features were more sensitive than structural ones, with eigenmode 17 demonstrating the greatest sensitivity (*d* in FRDA = 0.467). Correlations between geometric features and the standard clinical severity score (mFARS) are also reported in Supplementary Tables S14.

For benchmarking, we computed Cohen’s *d* for mFARS and conventional volumetric MRI measures (cerebellar volume, left/right superior cerebellar peduncle volume) in the TRACK-FA dataset. mFARS (*d* = 0.53) demonstrated sensitivity comparable to that of the top-performing predicted functional geometric feature. In contrast, volumetric measures were markedly less sensitive, with the largest absolute effect size in FRDA reaching only |*d*| = 0.11 and all between-group effect size differences remaining below 0.15. Overall, these results suggest that structural and predicted functional geometric features capture subtle longitudinal disease progression with greater sensitivity than traditional structural MRI biomarkers and may serve as objective, complementary measures to subjective clinical rating scales.

## Discussion

Our study provides proof-of-concept that cortical geometric eigenmodes--natural spatial patterns shaped by cortical geometry--encode subtype-specific signatures in HCAs. By leveraging geometric features derived from routinely available sMRI, our DL framework enabled differential diagnosis, prediction of fMRI-equivalent biomarkers, reliable approximation of individual-level task-activation patterns, and sensitive monitoring of disease progression. These biomarkers complement conventional sMRI by capturing multiscale cortical organisation, from local to network-level structure, and allow clinicians to access aspects of functional network disruption using routine sMRI, which is far more feasible in movement-disorder populations. Additionally, empirical functional geometric features further improved diagnostic precision, and structure-predicted functional biomarkers enhanced both diagnostic accuracy and sensitivity to longitudinal change. This framework establishes a scalable pathway to augment phenotypic characterisation, guide targeted genetic testing and potentially reduce diagnostic delay. Both structural and predicted functional geometric biomarkers provide objective, rater-independent measures of disease evolution that are more sensitive than conventional volumetric MRI features and demonstrate sensitivity comparable to established clinical rating scales.

Currently, MRI is primarily used as a complementary tool in a structured, rule-based, and semi-automated manner to guide differential diagnosis and support genetic testing.^45,46^ Existing structural MRI-based studies investigating automated diagnostic approaches for HCA subtypes are scarce, with only a few reporting accuracies comparable to our eigenmode-based classifier, though based on very small cohorts.^24,47^ Similarly, patient-control classification approaches for SCAs reported high accuracy, yet relied on small, single-site cohorts, lacked generalisability or external validation, and offered limited interpretability of features.^22,48^ In contrast, our framework enables automated differentiation among multiple HCA subtypes while demonstrating strong cross-cohort generalisability for patient-control classification using independent cohorts and rigorous cross-validation when external datasets were unavailable. After age-matching wherever required in the differential-diagnosis models to address this key confounder, the included cohorts exhibited comparable clinical severity, with mean scores within the moderate range for respective scales (Supplementary Tables S3), making severity-based confounding unlikely. Overall, the cross-cohort robustness observed here strengthens confidence that eigenmode-based biomarkers capture stable disease-related patterns and supports their potential for real-world deployment. This approach also underscores the potential of structure-based diagnostic strategies, paving the way for broader applications across other HCA subtypes and improved accuracy through integration of additional feature maps within the same pipeline, ML methodological refinement and larger, more heterogeneous training cohorts, especially for relatively difficult cases (such as distinguishing SCA subtypes).

Beyond diagnosis, we addressed another gap: predicting functional features from sMRI alone. A direct comparison with prior work was not possible, as only one previous study^31^ suggested that sMRI may be the most informative modality for predicting task-fMRI activation in healthy individuals using DL; however, it did not report any quantitative individual- or population-level prediction accuracy and has not been evaluated in disease cohorts. To our knowledge, this study represents the first demonstration of the feasibility of predicting fMRI equivalent features, leading to reliable approximation of individual activation maps from sMRI alone in both HCs and disease cohorts. Using 5-fold cross-validation, a rigorous approach for modest sample sizes, we achieved moderate-to-strong predictive performance (up to R² ≈ 0.6, correlation ≈ 0.8 for top modes) (Supplementary Tables S13). Unlike prior eigenmode-based work reporting >80% reconstruction accuracy for the task activation map at the population level,^34^ our estimates reflect individual-level reconstruction, explaining part of the performance gap. In healthy cohort, our findings support prior evidence that functional activities are primarily governed by low-order eigenmodes derived from diffusion MRI connectivity.^49^ In addition, the top predicted modes (Supplementary Tables S13) provide proof-of-concept that structure-function mapping persists across global and fine-grained scales, even in neurodegeneration, with signs of disrupted low-frequency mapping in the right hemisphere. Future studies should validate these results in larger samples and across disease stages.

We evaluated the utility of structure-predicted functional features using multiple approaches. In a large, independent cohort, these features achieved moderate diagnostic performance and integrating them with structural measures improved classification relative to structural markers alone, particularly in the left hemisphere, highlighting their practical utility when fMRI data are unavailable. In addition, several structural and predicted functional eigenmode features showed greater sensitivity to annual disease progression than conventional volumetric measures, with low-order functional mode 17 in the right hemisphere performing comparably to clinical scales, offering a novel, objective, surrogate biomarker. In support, a recent study in a separate cohort (IMAGE-FRDA) found no single volumetric feature qualified as a progression biomarker over two years; only inferior-posterior cerebellar volume (lobule VIII_IX) showed an opposite direction of progression between groups, with weak effect size (d ≈ 0.12) in FRDA.^26^ It is worth noting that although standard progression-biomarker criteria require only significant differences in rates of change between patient and control groups, without necessitating opposing directions of progression, we adopted this stricter criterion to improve the specificity and robustness of biomarker identification. Future research should examine the sensitivity of the geometric features across age groups, ambulation status and longer or shorter follow-up intervals.

To gain deeper insight into multiscale neurodegeneration patterns in FRDA, we analysed hemisphere-specific sensitivity of structural and predicted functional eigenmodes to disease progression. Geometric eigenmode analysis revealed distinct structural and functional network alterations over time, with sign reversals between FRDA and HCs, confirming disease-specific reorganisation. In the left hemisphere, structural low- to mid-order modes (e.g., 23, 52, 57; |*d*| ≈ 0.23 − 0.26) were most sensitive, suggesting disruption of large-scale organisation, such as cerebellar-cortical networks, while functional biomarkers peaked in higher-order modes (e.g., 124, 179; |*d*| ≈ 0.205), reflecting local desynchronisation. In the right hemisphere, structural disruption was greatest in mid- to higher-order modes (e.g., 69, 168, 184; |*d*| > 0.15), whereas functional alterations spanned low- and high-order modes (e.g., 17, 146, 156; |*d*| > 0.30), indicating combined global and local instability. Notably, very low-order modes (e.g., 2-10), which carry strong diagnostic signals, were not sensitive to annual progression, suggesting global patterns remain stable over short intervals or the changes are undetectable with current methods. Bilateral involvement of mid-order structural and high-order functional modes aligns with prior evidence from a dementia related study that higher-order eigenmode weights decay rapidly in disease while lower-order mode weights persist.^50^ These findings should inspire further research to explore similar patterns in other HCAs and identify which eigenmodes are disrupted in each condition. Moreover, prior work^51^ showed that eigenmode “uniqueness”, defined as individual deviations from group-average patterns, increases during adolescence and predicts mental health outcomes, with mid-frequency modes 37-49 strongly associated. This result underscores that eigenmode signatures could capture both disease-related and developmental variation. Given that some HCAs preferentially manifest in childhood or adolescence, disentangling these effects will be crucial. Future studies should leverage eigenmode uniqueness metrics and move toward individual-specific geometric eigenmodes as “brain fingerprints” rather than population templates, to enhance biological interpretability.

Finally, while our structure-to-function prediction framework shows promise, several limitations warrant attention. Selecting input features by linear correlation may oversimplify complex dependencies, and training independent models for each functional mode ignores inter-mode relationships. Future work should explore multivariate feature selection, multi-output models, and architectures leveraging spatial relationships (e.g., graph neural networks or surface-based CNNs) on larger samples. Incorporating additional geometric features (curvature, sulcal depth, gyrification) and complementary modalities (diffusion MRI, microstructural indices) could improve sensitivity but increase complexity, requiring trade-offs between accuracy and scalability. Mapping predictive modes to anatomical regions and validating models on independent datasets with ground-truth functional data will enhance interpretability and generalisability. A striking aspect of this study is achieving effective biomarkers for diagnosis and progression in FRDA using only the cerebral cortical analysis, despite the disease primarily affecting the cerebellum. However, this finding is consistent with prior work using the same passive-movement fMRI paradigm,^36^ which reported reduced somatosensory cortical activation in FRDA, supporting our eigenmode-based evidence that cortical geometry captures disease-related alterations. In addition, the somatomotor network emerges as one of the dominant functional territories underlying the HCs vs FRDA diagnostic functional eigenmodes. Spatial mapping of these modes also aligns with previous fMRI findings demonstrating altered task-related activation across somatomotor, limbic, and visual networks in FRDA.^3^ Consistent with this, the diagnostic structural modes in our study reveal cortical-thickness alterations in regions corresponding to these functional networks. Broadly across HCAs, previous studies reported cortical involvement at varying degrees: reduced gyrification in sensorimotor regions in FRDA,^52^ cortical grey matter loss in temporal regions in SCA1,^19^ and widespread cortical and subcortical atrophy in SCA3,^42,53^ likely contributing to the high diagnostic accuracy observed and the spatial dominance of specific networks in our differential classification models. The progression-sensitivity of functional modes also aligns with prior FRDA fMRI findings reporting cortical activity changes over time, although direct comparison is limited given different fMRI paradigms used.^3^ Nonetheless, future work integrating both cortical and cerebellar eigenmodes will be critical for brain-wide biomarkers and improved classification in challenging subtypes.

Overall, our results highlight a key insight that pathology and cortical architecture may co-evolve, with disease-related changes in cortical geometry and intrinsic structural and functional organisation mutually constraining one another over the course of HCA disease progression.

## Data Availability

Data are available upon reasonable request. Deidentified participant data underlying main results may be provided to researchers who contact the principal investigator of the respective dataset to establish a data-sharing agreement. Approval for data access will be granted on a case-by-case basis at the discretion of the principal investigator.

## Acknowledgements

The authors gratefully acknowledge the Friedreich’s Ataxia Research Alliance (FARA, USA) for funding this project through the Award for Innovative Mindset Grant Scheme (2024).

Full list of TRACK-FA Neuroimaging Consortium members: Professor Nellie GeorgiouKaristianis, Professor Louise A. Corben, Professor Eric F. Lock, Dr Helena Bujalka, Dr Isaac Adanyeguh, Dr Jonathan J. Cherry, Dr Manuela Corti, Professor Martin Delatycki, Dr Imis Dogan, Ms Jennifer Farmer, Professor Marcondes França, Dr Anthony S. Gabay, Professor William Gaetz, Professor Ian H. Harding, Professor Pierre-Gilles Henry, Dr James Joers, Ms Michelle A. Lax, Professor Christophe Lenglet, Mr Jiakun Li, Professor David Lynch, Dr Thomas Mareci, Professor Alberto R. M. Martinez, Professor Massimo Pandolfo, Dr Marina Papoutsi, Dr Richard G. Parker, Dr Myriam Rai, Professor Kathrin Reetz, Dr Thiago J. R. Rezende, Professor Timothy P. Roberts, Dr Sandro Romanzetti, Dr David A. Rudko, Dr Susmita Saha, Professor Jörg B. Schulz, Dr S. H. Subramony, Dr Veena G. Supramaniam.

The authors thank the research coordinators across all TRACK-FA sites for their assistance in participant recruitment and testing.

This work was supported by the MASSIVE HPC facility (www.massive.org.au).

